# Impact of Pharmacist-Led Deprescribing Interventions on Medication Related Outcomes Among Older Adults: A Systematic Review and Meta-analysis

**DOI:** 10.1101/2025.07.23.25331989

**Authors:** Zelalem Tilahun Tesfaye, Boressa Adugna Horsa, Malede Berihun Yismaw

**Author notes:** Corresponding author: Zelalem Tilahun Tesfaye.

## Abstract

**Background:** Older adults usually experience polypharmacy which increases their risk of adverse drug events, drug-drug interactions, and medication non-adherence. Clinical pharmacists, with specialized expertise in pharmacotherapy, are capable of engaging in deprescribing interventions aimed at reducing potentially inappropriate medications and overall medication burden. However, the overall impact of these pharmacist-led strategies remains unclear due to heterogeneity in study designs, settings, and outcomes.

**Objectives:** The aim of this study was to synthesize the available evidence on pharmacist-led deprescribing interventions on medication related outcomes among older adults.

**Methods:** We searched PubMed/MEDLINE, ScienceDirect, and the Cochrane Library for English language randomized controlled trials and high-quality nonrandomized studies published from January 2015 onward, comparing pharmacist-led deprescribing interventions to usual care in any setting (community, outpatient, hospital, or long-term care). Primary outcomes were mean change in total number of medications per patient and the proportion of patients achieving effective deprescribing (discontinuation of ≥1 PIM or ≥0.5 reduction in a drug burden index). Data were pooled using random effects models, and between-study heterogeneity was assessed with I^2. Subgroup analyses contrasted intensive versus less intensive pharmacist involvement.

**Results:** Seven studies (five RCTs, two nonrandomized) encompassing 3,607 older adults met inclusion criteria. The pooled mean difference (MD) in total medications at last follow-up favored intervention by -0.55 medications (95% CI: -2.17 to 1.07; I^2 = 83.1%), and the pooled risk ratio (RR) for effective deprescribing was 1.85 (95% CI: 0.63-5.45; I^2 = 73.5%), though neither reached statistical significance. In subgroup analyses, intensive interventions, characterized by comprehensive, in-person reviews, explicit deprescribing criteria, patient education, and direct physician outreach, yielded a significant reduction in medication count (MD -1.74; 95% CI: -2.86 to -0.62) and increased deprescribing (RR 3.55; 95% CI: 2.45-5.15). Less intensive approaches showed no clear benefit. Secondary outcomes indicated improvements in medication burden indices without increased adverse events.

**Conclusion:** Pharmacist-led deprescribing interventions, particularly when intensive and integrated within collaborative care models, appear promising for safely reducing polypharmacy in older adults. The high heterogeneity and limited reporting of clinical and patient-centered outcomes underscore the need for larger, standardized trials with longer follow-up to establish scalable, sustainable deprescribing practices.

**Protocol registration:** The protocol for this systematic review was registered in International Prospective Register of Systematic Reviews (PROSPERO identifier: CRD420251072072).

## Background

Older adults usually take complex medication regimens, often taking five or more prescription drugs concurrently (1). This polypharmacy increases the risk of adverse drug events, drug-drug interactions, and medication non-adherence (2, 3). Age-related changes in pharmacokinetics and pharmacodynamics, such as reduced renal clearance and altered body composition, make older people especially vulnerable to side effects and toxicity (4). In addition, cognitive impairment, sensory deficits, and multiple chronic conditions can make it hard for patients and caregivers to manage medications safely and effectively (5). As a result, hospitalizations related to falls, confusion, or organ dysfunction are more frequent in this population, and overall quality of life can decline as medication burdens mount (6, 7).

Various strategies have been developed to address these challenges. Clinical guidelines now emphasize regular medication reviews, deprescribing protocols, and use of assessment tools like the Beers Criteria or STOPP/START (8, 9). Multidisciplinary teams that include including physicians, nurses, and pharmacists may conduct structured medication reconciliations at care transitions or during routine visits. Educational programs for patients and training workshops for prescribers aim to improve awareness of potentially inappropriate medications (PIMs) and to encourage dose adjustments or safer alternatives (10, 11). While these efforts have shown promise in reducing certain risks, implementation remains inconsistent across settings, and outcomes vary widely depending on the model of care and the professionals involved.

With specialized training in pharmacotherapy and pharmaceutical care, clinical pharmacists can effectively identify PIMs, assess drug-drug and drug-disease interactions, monitor for adverse drug reactions, and recommend evidence-based adjustments. Pharmacist engagement in elderly care, in particular, is essential as older adults are often subjected to multiple concurrent medications, predisposing them to a higher risk of potential drug-related problems (12). Through medication therapy management sessions, pharmacists can engage older adults in shared decision-making, clarify dosing schedules, and monitor for adverse events, withdrawal symptoms as well as treatment failures (13, 14). In some settings, pharmacist-led clinics or collaborative practice agreements empower pharmacists to initiate or modify therapy under defined protocols (15, 16). By focusing on the medication use process from prescribing through monitoring, pharmacists can address gaps that might otherwise be missed in physician-only reviews (17, 18).

Despite growing interest in pharmacist-led deprescribing, the overall magnitude of benefit remains unclear. Individual studies report improvements in medication appropriateness, reductions in potentially harmful drug use, and, in some cases, lower rates of adverse events or health-care utilization (19–21). However, differences in study design, patient populations, intervention components, and measured outcomes make it hard to draw firm conclusions or establish best practices. Conducting a systematic review and meta-analysis will help synthesize the available evidence, quantify the effect size of pharmacist-led interventions, and identify factors that influence success. These insights can be valuable inputs for clinical guidelines, support wider adoption of pharmacist-driven models, and ultimately improve medication safety and quality of life for older people.

Although several systematic reviews and metanalyses have been published on the topic of deprescribing in older adults, the majority did not access the role of pharmacists in deprescribing (22–28). Systematic reviews that assessed pharmacist-led deprescribing focused on a specific setting which may limit the implications of their findings (29, 30). This systematic review was performed with the aim of accessing the impact of pharmacist-led deprescribing interventions among older adults across different settings.

## Methods

This systematic review and meta-analysis was conducted in accordance with the Preferred Reporting Items for Systematic Reviews and Meta-analyses (PRISMA) guidelines (31), and the protocol was registered in International Prospective Register of Systematic Reviews (PROSPERO identifier: CRD420251072072).

### Databases and Search Strategy

We searched multiple databases including PubMed/MEDLINE, ScienceDirect and Cochrane Library. In each search combinations of terms for pharmacists and deprescribing, older adults, polypharmacy/multiple medications, and study design were used (e.g. “(pharmacist OR “clinical pharmacist”) AND (deprescri* OR discontinu* OR cessation OR taper*) AND (older OR elderly OR geriatric OR “older adult” OR aged) AND (polypharm* OR “multiple medications” OR PIM OR “potentially inappropriate medication”) AND (randomized OR controlled OR trial OR cohort OR quasi)”. The references on the research articles retrieved from database searches were manually reviewed for additional articles that could be relevant for the review. The details of the search strategy are presented on Table 1 in the supplementary files.

**Table 1:**
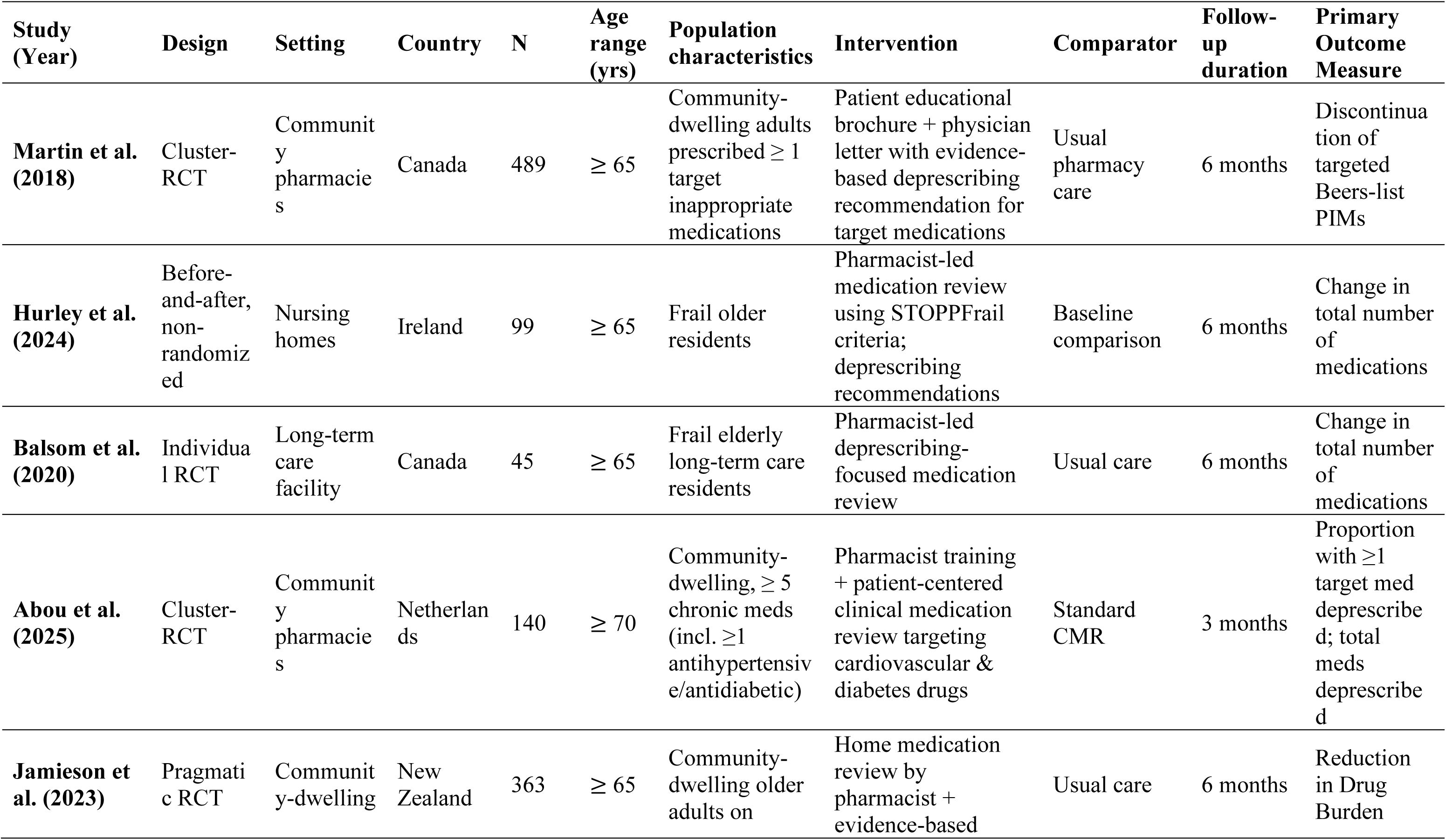
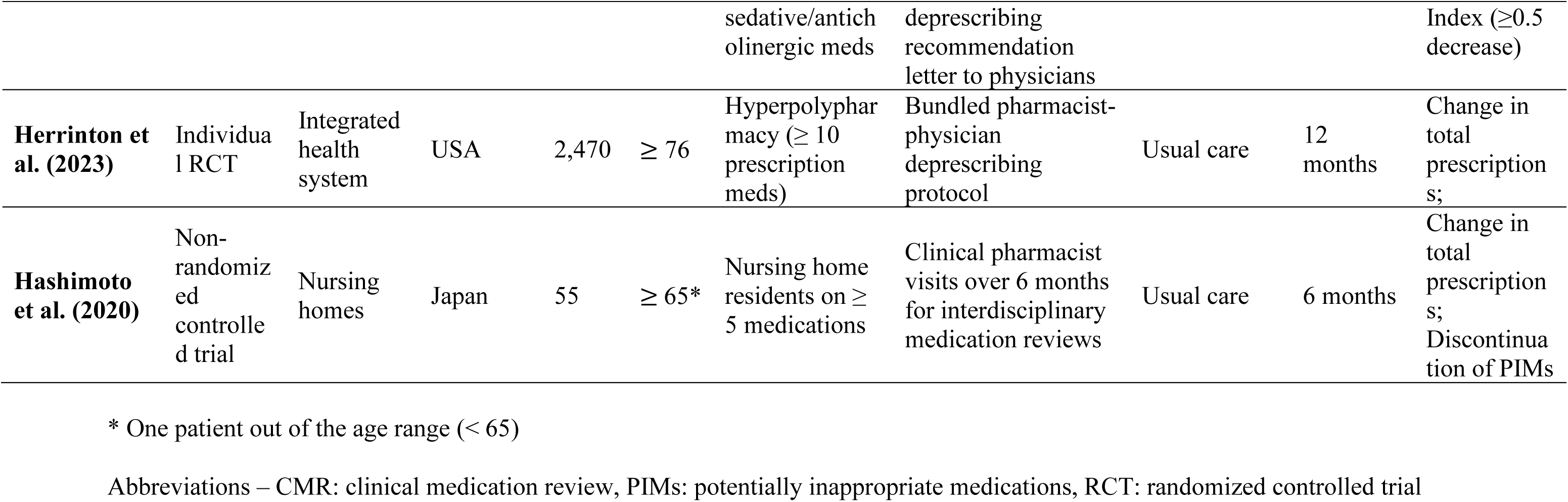
Characteristics of included studies.

| Study (Year) | Design | Setting | Country | N | Age range (yrs) | Population characteristics | Intervention | Comparator | Follow-up duration | Primary Outcome Measure |
| --- | --- | --- | --- | --- | --- | --- | --- | --- | --- | --- |
| **Martin et al. (2018)** | Cluster-RCT | Community pharmacies | Canada | 489 | $\geq 65$ | Community-dwelling adults prescribed $\geq 1$ target inappropriate medications | Patient educational brochure + physician letter with evidence-based deprescribing recommendation for target medications | Usual pharmacy care | 6 months | Discontinuation of targeted Beers-list PIMs |
| **Hurley et al. (2024)** | Before-and-after, non-randomized | Nursing homes | Ireland | 99 | $\geq 65$ | Frail older residents | Pharmacist-led medication review using STOPPFrail criteria; deprescribing recommendations | Baseline comparison | 6 months | Change in total number of medications |
| **Balsom et al. (2020)** | Individual RCT | Long-term care facility | Canada | 45 | $\geq 65$ | Frail elderly long-term care residents | Pharmacist-led deprescribing-focused medication review | Usual care | 6 months | Change in total number of medications |
| **Abou et al. (2025)** | Cluster-RCT | Community pharmacies | Netherlands | 140 | $\geq 70$ | Community-dwelling, $\geq 5$ chronic meds (incl. $\geq 1$ antihypertensive/antidiabetic) | Pharmacist training + patient-centered clinical medication review targeting cardiovascular & diabetes drugs | Standard CMR | 3 months | Proportion with $\geq 1$ target med deprescribed; total meds deprescribed |
| **Jamieson et al. (2023)** | Pragmatic RCT | Community-dwelling | New Zealand | 363 | $\geq 65$ | Community-dwelling older adults on | Home medication review by pharmacist + evidence-based | Usual care | 6 months | Reduction in Drug Burden |

| | | | | | | sedative/anticholinergic meds | deprescribing recommendation letter to physicians | | | Index ($\geq 0.5$ decrease) |
| --- | --- | --- | --- | --- | --- | --- | --- | --- | --- | --- |
| **Herrinton et al. (2023)** | Individual RCT | Integrated health system | USA | 2,470 | $\geq 76$ | Hyperpolypharmacy ($\geq 10$ prescription meds) | Bundled pharmacist-physician deprescribing protocol | Usual care | 12 months | Change in total prescriptions; |
| **Hashimoto et al. (2020)** | Non-randomized controlled trial | Nursing homes | Japan | 55 | $\geq 65^*$ | Nursing home residents on $\geq 5$ medications | Clinical pharmacist visits over 6 months for interdisciplinary medication reviews | Usual care | 6 months | Change in total prescriptions;<br>Discontinuation of PIMs |

\* One patient out of the age range ( $< 65$ )
Abbreviations – CMR: clinical medication review, PIMs: potentially inappropriate medications, RCT: randomized controlled trial

### Eligibility Criteria

We included studies on adults aged 65 or older taking regular medications that compared a pharmacist-led deprescribing intervention, such as structured medication review, education, or collaborative case conferencing, with usual care or standard medication management. Eligible designs were randomized controlled trials (RCTs), high-quality cohort studies, and quasi-experimental studies (for example, controlled before-and-after or interrupted time-series designs). To ensure relevance and consistency, we limited focused on peer-reviewed articles published in English from January 2015 onward, with no restriction on care setting (community-dwelling, outpatient, hospital, or nursing home). Studies were required to report at least one of our primary outcomes, change in total medication count or having received effective deprescribing intervention, identified by explicit criteria such as discontinuation of at least one PIM based on the Beers list, STOPP/START criteria, or reduction in drug burden indices.

### Data Extraction Procedures

Two reviewers independently extracted study characteristics and outcomes using a standardized, pilot-tested form to ensure consistency. For each study we recorded design and setting, sample size, participant’s mean/median age and female to male distribution, details of the pharmacist-led intervention and comparator, follow-up duration, and all relevant outcomes, including mean change in total number of medications (with measures of variability), number or proportion of patients with effective deprescribing interventions. We also noted which explicit PIM criteria each study used (for example, Beers, STOPP/START, or drug burden indices). Extracted data were cross-checked and any discrepancies were resolved through discussion; if needed, a third reviewer adjudicated.

### Quality Assessment

Risk of bias was evaluated for each study using the Cochrane Risk of Bias 2 (RoB 2) tool for randomized trials and the ROBINS-I tool for nonrandomized and quasi-experimental studies. Two reviewers independently assessed key domains, such as sequence generation, allocation concealment, blinding of participants and outcome assessors, completeness of outcome data, and selective reporting for RCTs, and confounding, participant selection, intervention classification, and measurement of outcomes for nonrandomized designs.

### Outcomes of Interest

Our primary outcomes were the change in total number of medications prescribed per patient, and the proportion of patients with effective deprescribing interventions (discontinuation of least one PIM or ≥0.5 reduction in drug burden index), as these directly reflect the impact of pharmacist-led deprescribing on polypharmacy and inappropriate prescribing. We included only studies that reported mean (or median) change in medication count with measures of variability or provided the number or percentage of patients discontinuing or remaining on PIMs, based on explicit criteria such as Beers, STOPP/START, or drug burden indices. Secondary outcomes of interest, when reported, included adverse drug events, hospital admissions, falls, measures of regimen complexity, quality-of-life indicators, process implementation, and cost effectiveness, allowing us to explore potential benefits or harms beyond medication reduction.

## Results

### Study Selection

After combining all records and removing duplicates, 1,035 unique records were screened by title and abstract. From these, 982 were excluded for reasons including wrong population, wrong intervention (not pharmacist-led deprescribing), or wrong outcomes (no medication count or deprescribing rate indicator). The remaining 53 full-text articles were reviewed in detail. Of these, 46 were excluded for reasons such as: not a controlled design (e.g. reviews, protocols), outcome of interest not measured, or studies not with non-pharmacist interventions. Finally, 7 studies met all inclusion criteria and were selected for the systematic review. The PRISMA flow of identification and screening is summarized on Figure 1.

**Figure 1:**
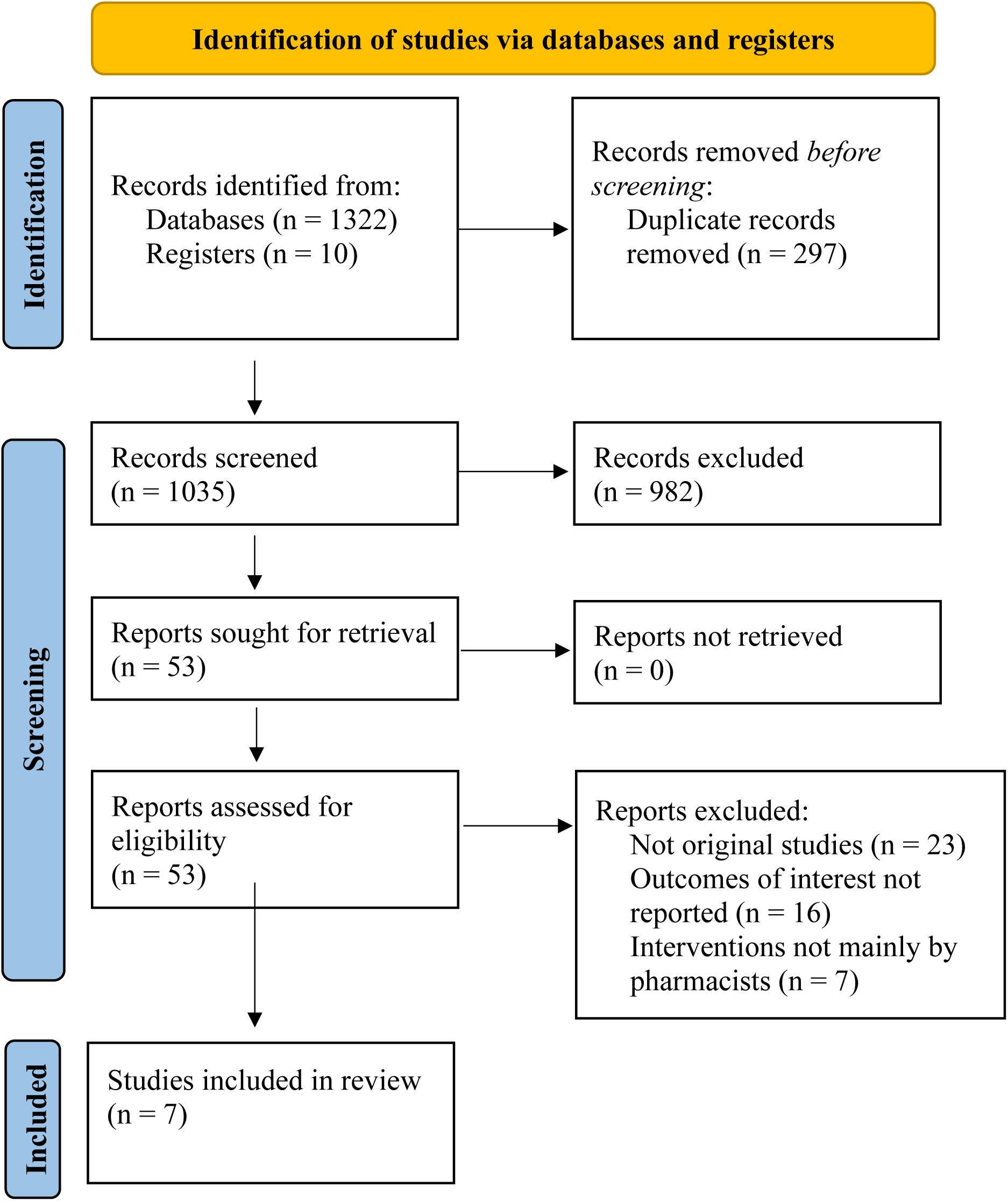
Flow diagram of the process of selection of included studies.

### Included Studies

Seven studies (5 RCTs and 2 non-randomized studies) met inclusion criteria. All studies are published in peer-reviewed journals from 2018 to 2025. Three studies were conducted in long-term care facilities, and four were conducted in community settings. A total of 3607 individuals were included in the studies. All included studies targeted older adults on prescription medications, comparing a pharmacist-led deprescribing strategy to usual care.

Martin et al. (32) reported the D-PRESCRIBE trial, a cluster RCT in community pharmacies in Quebec. Sixty-nine pharmacies were randomized (34 intervention, 35 control). Eligible patients were community-dwelling adults aged ≥65 years prescribed at least one of four target inappropriate medications (sedative-hypnotics, first-generation antihistamines, glyburide, or non-steroidal anti-inflammatory drugs (NSAIDs)). In the intervention arm, pharmacists delivered a consumer-focused educational brochure to each patient and concurrently sent an evidence-based deprescribing recommendation letter to the patient’s physician. Control pharmacies provided usual care. The primary outcome was discontinuation of the target medication at 6 months, confirmed via pharmacy refill records.

Hurley et al. (33) evaluated a pharmacist-led deprescribing program using the STOPPFrail criteria in nursing homes. Ninety-nine frail older residents across six nursing homes were included. A clinical pharmacist reviewed each patient’s medications using STOPPFrail and generated deprescribing recommendations for potentially burdensome or inappropriate medications. These were presented to each resident’s general practitioner for potential implementation. Outcomes measured at baseline, immediately post-review, and at 6 months included total medication count, monthly medication cost, drug burden index (DBI), anticholinergic cognitive burden (ACB) score, a medication appropriateness index (MMAI), and health events such as falls and hospitalizations.

Balsom et al. (34) reported a single-center randomized trial in a long-term care home. Forty-five frail elderly residents were randomized to a pharmacist-led intervention versus usual care. The pharmacist performed a comprehensive medication review for intervention patients, identifying potentially inappropriate or unnecessary drugs and making recommendations to the care team. The control group continued with standard care. The study assessed change in medication count at 3 and 6 months as the primary outcome.

Abou et al. (35) conducted the LeMON study, a cluster RCT in community pharmacies. Twenty community pharmacists from a nationwide chain were randomized to receive deprescribing-focused training or to usual care. The trial enrolled older primary-care patients aged 70 years or older with polypharmacy (≥5 chronic medications, including at least one antihypertensive or antidiabetic) and good recent blood pressure or HbA1c control. Pharmacists in the intervention arm received training to perform a patient-centered clinical medication review (CMR) targeting cardiovascular and diabetes drugs and then conducted CMRs with patients (in-person or by phone), incorporating shared decision-making. Twenty-one community pharmacists completed CMRs for 140 patients (71 intervention, 69 control). The main outcome was the proportion of patients with one or more cardiovascular or antidiabetic medications deprescribed within 3 months of the CMR.

Jamieson et al. (36) conducted a community-based cluster RCT stratified by frailty. Using interRAI assessment registries, they enrolled 363 older adults living in the community and taking sedative or anticholinergic medications. Participants were randomized to usual care or a pharmacist-led deprescribing intervention. The intervention consisted of pharmacist-conducted home medication reviews and evidence-based deprescribing recommendations sent to the patient’s general practitioner, specifically targeting sedative-hypnotic and anticholinergic drugs. The primary outcome was the proportion of participants achieving a reduction of ≥0.5 in DBI within 6 months.

Herrinton et al. (37) performed a large pragmatic RCT in an integrated care system. A total of 2,470 patients aged 76 years or older, each using ≥10 prescription medications, were identified in primary care clinics and randomized 1:1. Patients in the intervention arm received a bundled pharmacist-led deprescribing program that involved a physician-pharmacist collaborative medication management process with shared decision-making and deprescribing protocols administered primarily by telephone over multiple encounters for up to 180 days. Usual care patients continued with standard medication management. The trial’s coprimary outcomes were the change in total medication count and the prevalence of geriatric syndromes (falls, cognitive impairment, urinary incontinence, and pain) comparing pre-randomization to one-year post-randomization.

Hashimoto et al. (38) conducted a non-randomized controlled trial in four special elderly nursing homes. Two facilities were assigned to the pharmacist-led intervention and two to usual care. Older nursing home residents, all taking ≥5 medications, were eligible; data from 28 intervention and 27 control residents were analyzed. A clinical pharmacist visited each intervention home over 6 months to perform interdisciplinary medication reviews, identify drug-therapy problems, and advise staff and physicians. The control homes received standard care without pharmacist review. Primary outcomes included the incidence of PIMs use and falls.

The major characteristics of the included studies are summarized on Table 1.

### Summary of Study Findings

Across the seven included studies, pharmacist-led deprescribing interventions produced mixed results on our outcomes interest. In trials measuring the mean difference in total medications, two studies showed significant reductions with pharmacist input, while one large trial found no effect. In a Canadian nursing home RCT, Balsom et al. (34) found the intervention group took an average 2.68 fewer at 3 months and 2.88 fewer at 6 months (p<0.02). Similarly, Hurley et al. (33) reported a substantial drop in mean medication count: from 16.0 at baseline to 14.6 immediately after the pharmacist review (p<0.001), with a partial rebound to 15.4 at 6 months. In contrast, Herrinton et al. (37) and Hashimoto et al. (38) reported only a slight decline in medications dispensed in both groups and no significant difference between arms. (Table 2).

**Table 2:**
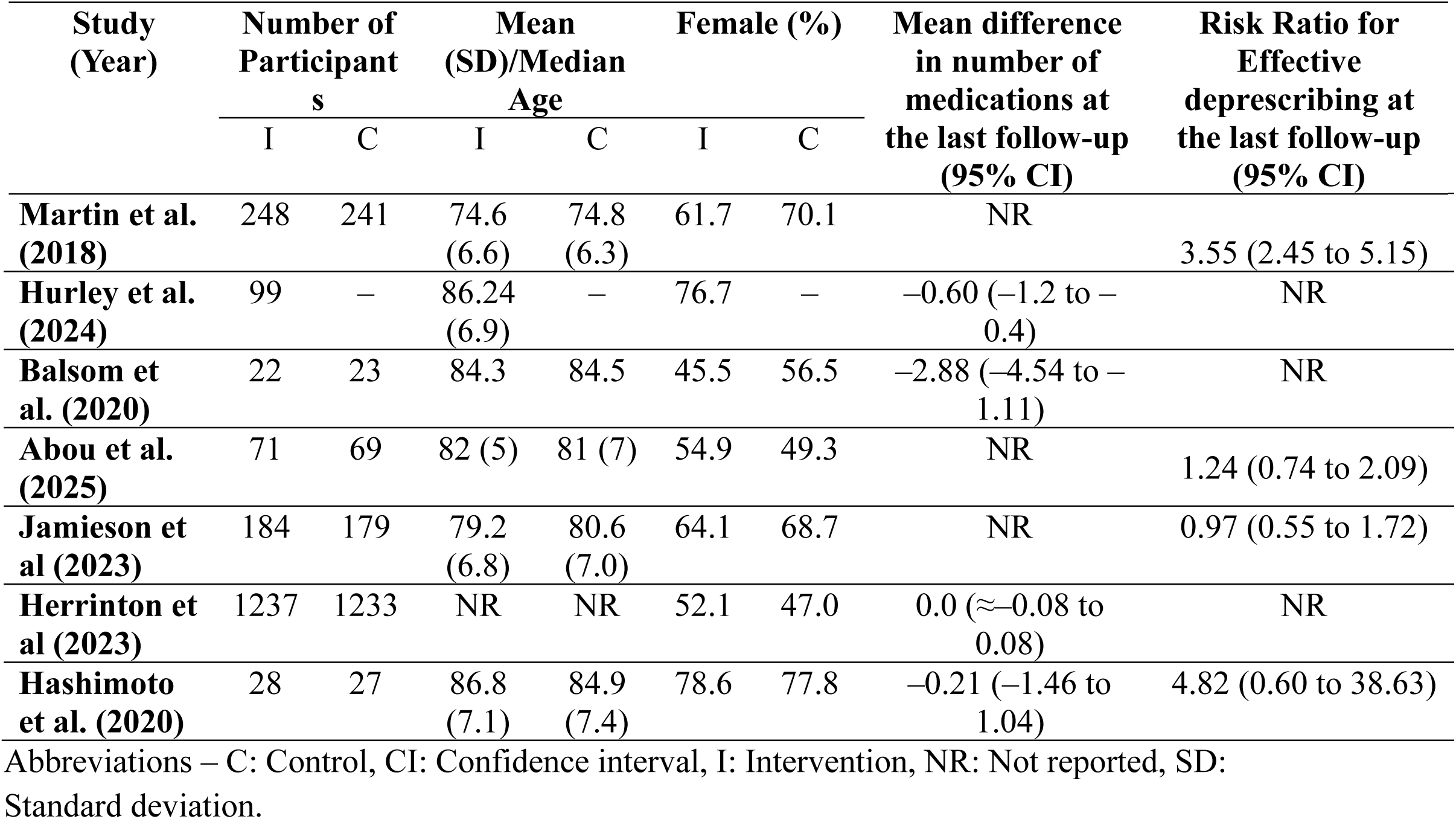
Summary of study results.

Results for the percentage of patients with effective deprescribing (defined as stopping ≥1 PIM, or DBI reduction ≥0.5) were also heterogeneous. Martin et al. (32) found a large effect: 43% of intervention patients had discontinued at least one targeted inappropriate drug by 6 months versus only 12% of controls. This trial showed significantly more patients ‘deprescribed’ under the pharmacist led educational intervention (RR: 3.55, 95% CI: 2.45-5.15) (32). By comparison, Jamieson et al. (36) found essentially no benefit: 12.2% of the intervention and 12.7% of control patients achieved a ≥0.5 DBI reduction (RR: 0.97, 95% CI: 0.55 to 1.72). Similarly, Abou et al.’s LeMON found only a nonsignificant trend: 51% of intervention patients vs 36% of controls had any medication discontinued (RR: 1.24, 95% CI: 0.74 to 2.09), and 32% vs 26% stopped a cardiovascular/diabetes drug (p=0.413) (35). Hashimoto et al. (38) reported a non-significant reduction in total number of medications in the intervention arm (mean difference: –0.21, 95% CI:–1.46 to 1.04), and 17.9% of intervention patients vs 3.7% of controls had any PIM reduced (p=0.094) (Table 2).

### Risk of Bias Assessment

Risk of bias was assessed by using Cochrane RoB 2. Key concerns included lack of blinding (all trials were open-label) and potential selection bias in the small studies (Figure 2). The details of reviewers’ judgments of individual studies risk bias is available in the supplementary files.

**Figure 2:**
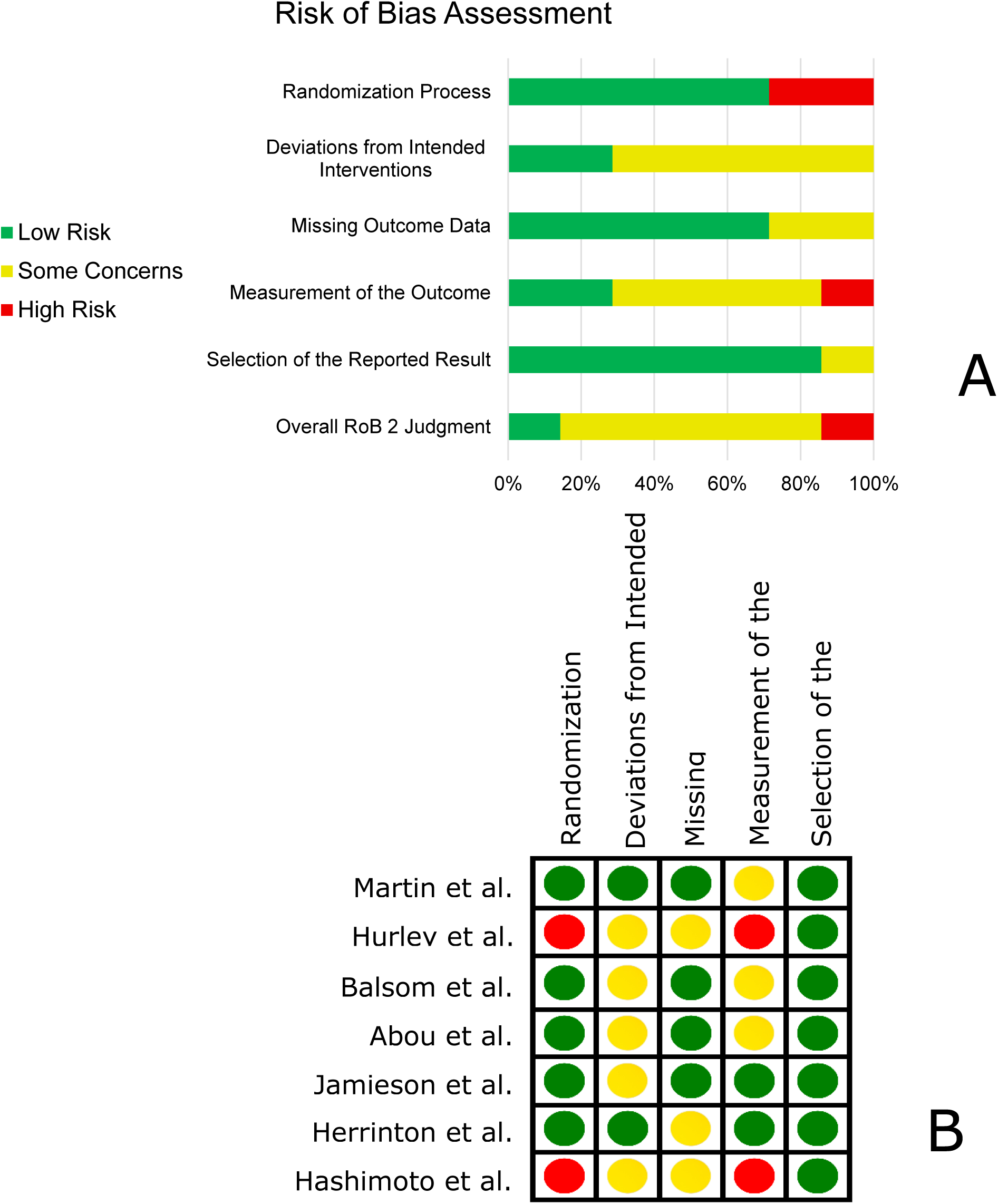
Risk of bias assessment; A: summary of risk of bias of all included studies, B: Individual risk of bias assessment of included studies

**Figure 3:**
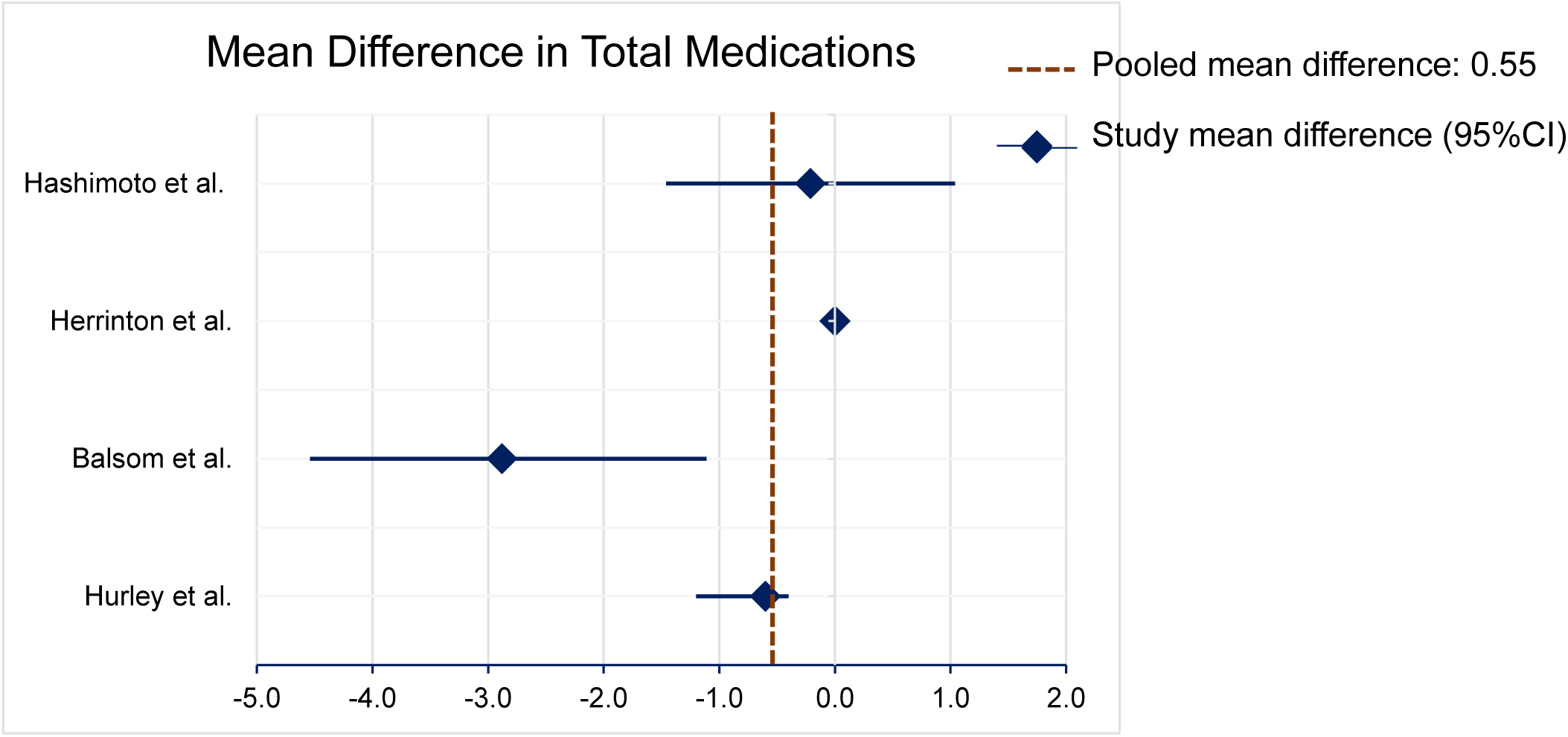
Forest plot of the pooled mean difference in number of medications between pharmacist-led deprescribing interventions and usual care.

**Figure 4:**
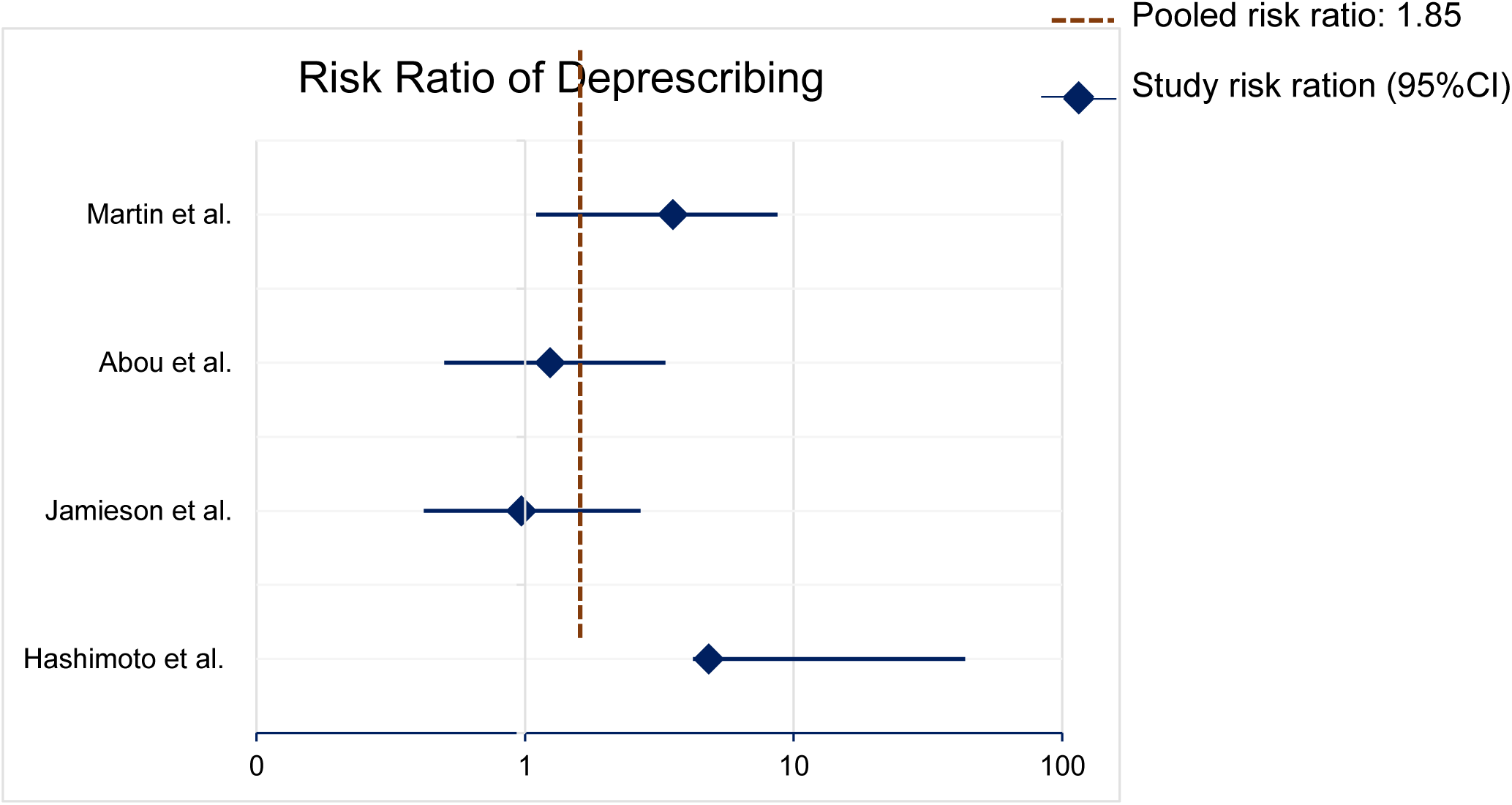
Forest plot of the pooled risk ratio for effective deprescribing at follow-up after pharmacist-led deprescribing vs. usual care.

### Random Effects Meta-analysis

The result of random effects meta-analyses revealed that the pooled mean difference in total medications between intervention and control at the last follow-up is −0.55 (95% CI −2.17, 1.07). Between-study variance (τ²) among the included studies was 2.35 and the heterogeneity (I²) was 83.1%. The pooled RR of effective deprescribing interventions was 1.85 indicating higher odds of deprescribing in the intervention arm despite not statistically significant (95% CI: 0.63–5.45, I^2^ : 73.5%).

### Subgroup Analysis

Subgroup analysis was performed based on the level of pharmacist involvement. Thus, Balsom et al. (explicitly deprescribing-focused comprehensive in-person medication reviews) (34), Hurley et al. (systematic use of STOPPFrail criteria, a specialized tool for deprescribing in end-of-life or severely frail populations) (33), and Martin et al. (use of personalized educational materials to patients, and evidence based recommendation for physicians, trained pharmacists) (32), were classified as studies with intensive pharmacist involvement while the rest were designated to have less intensive pharmacist involvement as they involved indirect contact with prescribers, lesser authority to pharmacists and/or infrequent reviews.

As a result, intensive pharmacist involvement is associated with a larger and statistically significant reduction in medication number (Mean difference: −1.74), whereas less intensive interventions show no clear effect. Regarding the rate of effective deprescribing, the single intensive-intervention study (Martin et al. (32)) reported a strong effect (RR:3.55) which was statistically significant, while among the less intensive group, the pooled RR (1.17) was not significant. Studies in each subgroup showed low level of heterogeneity (Table 3).

**Table 3:** Sub-group analysis based on the level of pharmacist involvement in deprescribing interventions.

| Group | Pooled Variable | Number of Studies | $\tau^2$ | I<sup>2</sup> (%) | Pooled Value (95% CI) |
| --- | --- | --- | --- | --- | --- |
| Intensive pharmacist involvement | MD in Number of Medications | 2 | 0.00 | — | -1.74 (-2.86, -0.62)* |
|  | RR for Effective Deprescribing | 1 | — | — | 3.55 (2.45, 5.15)* |
| Less intensive pharmacist involvement | MD in Number of Medications | 2 | 0.20 | 45.0 | -0.10 (-0.98, 0.78) |
|  | RR for Effective Deprescribing | 3 | 0.017 | 10.7 | 1.17 (0.77, 1.78) |

Abbreviations – CI: Confidence interval, MD: Mean difference, RR: Risk ratio
Symbols – $\tau^2$ : Between-study variance, I<sup>2</sup>: heterogeneity

### Summary of Secondary Outcomes

Pharmacist-led deprescribing interventions were also evaluated on several patient, process and cost related endpoints including medication burden indices and appropriateness scores, clinical and geriatric events, implementation fidelity, economic/resource use measures, and patient-reported outcomes. Findings are summarized below in a narrative synthesis.

### Medication Burden and Appropriateness Indices

Hurley et al. (33) reported significant immediate reductions in DBI, ACB score, and MMAI following the STOPPFrail-based review; these improvements persisted at six months.

### Clinical Events and Geriatric Syndromes

Several trials assessed hard clinical endpoints. Hashimoto et al. (38) demonstrated a markedly lower fall rate at six months in nursing homes receiving pharmacist-led reviews (3.6% versus 22.2% of residents). Herrinton et al. (37) included a composite of falls, cognitive impairment, urinary incontinence, and pain as co-primary outcomes but reported no significant between-group differences at one year among the hyperpolypharmacy population. Hurley et al. (33) confirmed that deprescribing did not increase adverse events; and falls, emergency visits, or hospitalizations remained stable post-intervention.

### Implementation and Process Measures

Adoption of pharmacist recommendations was consistently high. Balsom et al. (34)achieved an 85.1% uptake of deprescribing suggestions by the nursing-home care team, while Hurley et al. (33) saw 58.7% of clinically relevant recommendations accepted by general practitioners. Martin et al.’s D-PRESCRIBE trial verified true discontinuation via pharmacy refill data, underscoring strong implementation and measurement of deprescribing actions (32).

### Economic and Resource Use Outcomes

Only Hurley et al. (33) formally assessed medication costs, finding a modest but statistically significant reduction in average monthly drug expenditures immediately after the pharmacist review. None of the other studies conducted full cost-effectiveness analyses or reported broader health-care utilization metrics beyond hospitalization and emergency visits.

### Quality of life and patient-reported outcomes

Quality-of-life measures were infrequently reported. Hurley et al. (33) used a standardized instrument and found no significant change in residents’ quality of life at six months post-deprescribing. Other trials did not include structured patient-reported outcome instruments, limiting insight into the experiential impact of deprescribing on older adults.

## Discussion

This systematic review and metanalysis explored the impact of pharmacists educational and clinical interventions on older adults who are on regular medications. We assessed medication related outcomes as primary outcomes and a variety of patient, process and cost-related outcomes as secondary outcomes. Random effects metanalysis and subgroup analysis of the primary outcomes was performed to quantize the overall effect of pharmacist interventions on deprescribing.

The pooled estimate for mean difference in total medications at last follow-up favored pharmacist-led interventions by –0.55 medications (95% CI –2.17 to 1.07), but this effect did not reach statistical significance and was marked by substantial heterogeneity (I² = 83.1%). Similar level of reduction in number of medications has been reported by an non-provider-specific metanalysis (25). While two trials (Balsom et al. and Hurley et al.) (33, 34) demonstrated significant reductions of mean total medications in intervention arms, other two studies (Herrinton et al. and Hashimoto et al.) (37, 38) showed minimal change and no between-group difference. These mixed results mirror a prior metanalysis of general deprescribing interventions, which have similarly reported modest average reductions mean number of medications yet wide variability across studies (22).

Across four studies reporting dichotomous deprescribing outcomes, the pooled risk ratio for effective deprescribing (at least one PIM discontinued or ≥0.5 DBI reduction) was 1.85 (95% CI 0.63–5.45; I² = 73.5%), indicating a trend favoring intervention but lacking statistical significance. Notably, the D-PRESCRIBE trial by Martin et al. (32) reported a robust effect (RR: 3.55), whereas Jamieson et al. (36) and Abou et al. (35) observed minimal or nonsignificant benefits. This pattern underscores that while pharmacist involvement can drive deprescribing when interventions are directive and patient-centered, more passive or less intensive models may fail to translate into meaningful discontinuations (39, 40).

When stratified by intervention intensity, intensive pharmacist-led strategies (comprehensive in-person reviews, use of explicit tools, patient education with physician outreach) yielded a significant mean reduction of –1.74 medications (95% CI –2.86 to –0.62), compared with no clear effect in less intensive approaches (MD –0.10; 95% CI –0.98 to 0.78). Similarly, only the intensive subgroup demonstrated a significant increase in deprescribing (RR 3.55). These findings align with implementation science literature emphasizing that multifaceted, proactive interventions with direct patient engagement and provider collaboration are more effective than passive recommendations alone (41–43).

Pharmacist-driven reviews produced consistent short-term improvements in medication burden indices (DBI, ACB, MMAI), as seen in Hurley et al. (33), without increasing adverse events or geriatric syndromes. Clinical outcomes (falls, hospitalizations, cognitive impairment) were variably reported; one study showed reduced falls (38), while a larger trials found no significant differences (37). Economic data are scant but suggest modest monthly cost savings post-review (33). Quality-of-life and patient-reported outcomes were infrequently assessed, limiting conclusions about the experiential impact of deprescribing.

Our findings suggest that intensive, structured pharmacist-led deprescribing, characterized by comprehensive medication reviews using explicit criteria, direct patient education, and proactive communication with prescribers, can meaningfully reduce polypharmacy in older adults. To translate this into practice, health systems should expand collaborative practice agreements that empower pharmacists to initiate deprescribing protocols under physician oversight and invest in pharmacist training on validated tools and shared decision-making techniques. Embedding pharmacists within multidisciplinary care teams, particularly during high-risk transitions such as hospital discharge or nursing-home admission, may further enhance medication safety, lower medication-related costs, and improve patient satisfaction, all while ensuring regular monitoring for unintended harms (44–47).

## Conclusion

Our metanalysis indicates that pharmacist-led deprescribing interventions, particularly those that are intensive, patient-centered, and integrated within collaborative care models, show promise for safely reducing polypharmacy among older adults. Variability in study designs and outcomes underscores the need for larger, thoroughly designed trials with standardized protocols, longer follow-up, and comprehensive evaluations of clinical, economic, and patient-reported outcomes to establish scalable, sustainable deprescribing practices across diverse healthcare settings.

## Data Availability

All data produced in the present work are contained in the manuscript

