## Supplementary materials for "Impact of Pharmacist-Led Deprescribing Interventions on Medication Related Outcomes Among Older Adults: A Systematic Review and Meta-analysis"

**Supplementary Table 1:** Search Strategies for different Databases

| **Database** | **Search String** |
| --- | --- |
| PubMed/MEDLINE  (815 Results) | “(pharmacist OR “clinical pharmacist”) AND (deprescri* OR discontinu* OR cessation OR taper*) AND (older OR elderly OR geriatric OR “older adult” OR aged) AND (polypharm* OR “multiple medications” OR PIM OR “potentially inappropriate medication”) AND (randomized OR controlled OR trial OR cohort OR quasi)” |
| Cochrane Library  (10 Results) | Pharmacy led deprescribing |
| Science Direct  (507 Results) | Pharmacy led deprescribing |

**Supplementary Table 2:** Assessment of risk of bias using RoB 2 (Randomized Trials)

| **Study (Year)** | **Randomization Process** | **Deviations from Intended Interventions** | **Missing Outcome Data** | **Measurement of the Outcome** | **Selection of the Reported Result** | **Overall RoB 2 Judgment** |
| --- | --- | --- | --- | --- | --- | --- |
| **Abou et al. (2025)** (cluster‐RCT) | Low risk (clusters randomly allocated; allocation sequence concealed at cluster level) | Some concerns (providers knew allocation; no individual‐level blinding; possibility of performance bias) | Low risk (> 95 % follow-up reported for primary outcome; attrition balanced) | Some concerns (outcome assessors not formally blinded; primary endpoint = medication count, relatively objective) | Low risk (protocol published; prespecified outcomes all reported) | **Some concerns** |
| **Balsom et al. (2020)** (individual RCT) | Low risk (central randomization; adequate allocation concealment) | Some concerns (no blinding of pharmacists or care staff; potential behavior change in control group) | Low risk (> 90 % completion; reasons for missing data balanced) | Some concerns (outcome = number of medications; abstraction from charts by unblinded staff) | Low risk (trial registry and protocol matched published outcomes) | **Some concerns** |
| **Herrinton et al. (2023)** (individual RCT) | Low risk (computer‐generated randomization; allocation concealed) | Low risk (intention‐to‐treat analysis; minimal deviations) | Some concerns (~10 % lost to follow-up for primary endpoint; reasons appear unrelated to intervention) | Low risk (primary outcome = prescription fills, extracted from electronic health records) | Low risk (all prespecified outcomes reported in main publication) | **Low risk** |
| **Jamieson et al. (2023)** (individual RCT) | Low risk (1:1 randomization via central service; concealment adequate) | Some concerns (no masking of participants/providers; possibility of co‐interventions) | Low risk (> 95 % follow-up; missing data imputed as prespecified) | Low risk (blinded adjudication of deprescribing events) | Low risk (pre-registered trial with complete outcome reporting) | **Some concerns** |
| **Martin et al. (2018)** (individual RCT) | Low risk (randomization list generated offsite; sealed envelopes used) | Low risk (participants and pharmacists aware of group, but intention‐to‐treat maintained; minimal cross-overs) | Low risk (> 98 % follow-up for primary outcome) | Some concerns (assessment of “appropriate prescription” by pharmacists not fully blinded) | Low risk (primary and secondary endpoints reported as in protocol) | **Some concerns** |

**Supplementary Table 3:** Assessment of risk of bias using ROBINS-I (Non-Randomized Studies)

| **Study (Year)** | **Bias Due to Confounding** | **Bias in Selection of Participants** | **Bias in Classification of Interventions** | **Bias Due to Deviations from Intended Interventions** | **Bias Due to Missing Data** | **Bias in Measurement of Outcomes** | **Bias in Selection of the Reported Result** | **Overall ROBINS-I Judgment** |
| --- | --- | --- | --- | --- | --- | --- | --- | --- |
| **Hashimoto et al. (2020)** (non-randomized controlled) | Moderate (no detailed adjustment for baseline frailty or comorbidities; possible selection of less-complex cases into intervention) | Moderate (participants self-selected or referred by providers—possible systematic differences) | Low risk (intervention clearly defined; classification unlikely to be in error) | Moderate (intervention delivered by pharmacists vs “usual care” could entail ancillary support; some co-interventions not documented) | Some concerns (~15 % attrition in control vs 10 % in intervention; reasons partially described) | Some concerns (primary outcome = medication count via chart review by non-blinded staff) | Low risk (all prespecified outcomes reported; no evidence of selective reporting) | **Moderate** |
| **Hurley et al. (2024)** (non-randomized intervention) | Serious (no randomization; high likelihood that participants opting in differed systematically in baseline polypharmacy risk and motivation) | Serious (recruitment via provider referral; control group less clearly defined) | Low risk (intervention defined by application of STOPP/START tool; classification unlikely misassigned) | Moderate (lack of blinding; pharmacists and providers aware; possible differential co-interventions) | Moderate (~20 % missing follow-up medication data; missingness not fully explored) | Serious (outcome = self-reported falls and chart‐derived prescriptions, assessed by unblinded staff) | Some concerns (trial protocol not publicly available; secondary endpoints not fully described) | **Serious** |
